# Relationship between macronutrients, dietary components, and objective sleep variables measured by smartphone applications

**DOI:** 10.1101/2024.07.25.24311028

**Authors:** Jaehoon Seol, Masao Iwagami, Megane Kayamare, Masashi Yanagisawa

## Abstract

**Background:** Few studies have used daily data from objective applications to explore macronutrient interactions in sleep and nutrition research.

**Objective:** This cross-sectional study examined the relationships between macronutrients, dietary components, and sleep parameters, considering their interdependencies.

**Methods:** Data from 4,825 users of sleep and nutrition apps for at least 7 days were analyzed. Multivariable regression analysis investigated associations between quartiles of macronutrients (protein, carbohydrate, and total fat, including saturated, monounsaturated, and polyunsaturated fats), dietary components (sodium, potassium, dietary fiber, and sodium-to-potassium ratio), and sleep variables (total sleep time [TST], sleep latency [SL], and % wakefulness after sleep onset [%WASO]). Nutrients were divided into quartiles, with the lowest intake group as the reference. Compositional data analysis accounted for interdependencies among macronutrients. Analyses were adjusted for age, sex, and body mass index (BMI).

**Results:** Higher protein intake was associated with longer TST in the 3rd (B = 0.17, 95% CI = 0.09, 0.26) and 4th quartiles (B = 0.18, 95% CI = 0.09, 0.27). Higher total fat intake was linked to shorter TST in the 3rd (B = -0.11, 95% CI = -0.20, -0.27) and 4th quartiles (B = -0.16, 95% CI = -0.25, -0.07). Higher carbohydrate intake was associated with shorter %WASO in the 3rd (B = -0.82, 95% CI = -1.37, -0.26) and 4th quartiles (B = -0.57, 95% CI = -1.13, -0.01), while higher total fat intake was linked to longer %WASO in the 4th quartile (B = 0.62, 95% CI = 0.06, 1.18). Higher dietary fiber intake was consistently associated with longer TST in the 3rd (B = 0.11, 95% CI = 0.02, 0.19) and 4th quartiles (B = 0.18, 95% CI = 0.09, 0.26), shorter SL in the 2nd (B = -1.71, 95% CI = -2.66, -0.76), 3rd (B = -2.23, 95% CI = -3.19, -1.27), and 4th quartiles (B = -2.30, 95% CI = -3.27, -1.33), and shorter %WASO in the 2nd (B = -1.06, 95% CI = -1.61, -0.51), 3rd (B = -1.04, 95% CI = -1.59, -0.48), and 4th quartiles (B = -1.05, 95% CI = -1.61, -0.48). A higher sodium-to-potassium ratio was linked to shorter TST in the 3rd (B = -0.11, 95% CI = -0.20, -0.02) and 4th quartiles (B = -0.19, 95% CI = -0.28, -0.10), longer SL in the 2nd (B = 1.03, 95% CI = 0.08, 1.98) and 4th quartiles (B = 1.50, 95% CI = 0.53, 2.47), and longer %WASO in the 4th quartile (B = 0.71, 95% CI = 0.15, 1.28). Compositional data analysis, involving 6% changes in macronutrient proportions, showed higher protein intake correlated with longer TST (B = 0.27, 95% CI = 0.18, 0.35), while more monounsaturated fats were linked to longer SL (B = 4.64, 95% CI = 1.93, 7.34) and %WASO (B = 2.21, 95% CI = 0.63, 3.78). Higher polyunsaturated fat intake correlated with shorter TST (B = -0.22, 95% CI = -0.39, -0.05), shorter SL (B = -4.72, 95% CI = -6.58, -2.86), and shorter %WASO (B = -2.00, 95% CI = -3.08, -0.92).

**Conclusions:** These findings highlight the intricate relationships between dietary factors and sleep outcomes. Prospective studies are warranted to determine whether dietary interventions result in positive sleep outcomes.

## Introduction

Dietary intake patterns vary widely depending on the lifestyle. Brain imaging reveals that these patterns are associated with mental health, cognitive function, blood and metabolic biomarkers, overall brain structure, and white matter integrity [1]. Diet and sleep are closely linked [2]. A systematic review partially revealed that overeating, especially meals high in fat, can extend SL, increase nocturnal awakenings, and ultimately reduce TST [3].

Dietary intake and sleep are known to have a bidirectional relationship [2]. The intake of healthy foods is associated with improved sleep quality, whereas consuming processed foods and foods high in free sugars has been shown to deteriorate sleep quality [4]. Several hypotheses have been proposed to explain this relationship [5–7]. One hypothesis suggests that an increased dietary intake of tryptophan can promote the production of serotonin and melatonin, potentially leading to improved sleep quality [5]. Additionally, sleep deprivation can decrease leptin levels and increase ghrelin levels, a hormone that stimulates appetite, and may trigger overeating, especially at night, creating a negative spiral [5–7].

The importance of assessment methods in examining the relationship between the two has been emphasized in numerous studies [2,3]. Various traditional methods are available to assess dietary intake, including dietary diaries, 24-h recalls, food frequency questionnaires, and screening tools, which are traditional approaches [8]. In recent years, digital applications, such as smartphones, have been developed to record dietary intake in real time, thereby minimizing recall bias [8]. Furthermore, systems have been introduced to eliminate recording and subjective biases, in which nutritional components are automatically calculated by registering menus or scanning the barcodes of commercially available products [9]. Compared to dietary assessments, evaluating sleep is more challenging. However, in recent years, polysomnography, often considered the gold standard for sleep assessment, has been adapted from novel methods using portable EEG devices and accelerometers to more accessible methods, such as smartphone accelerometers and tappigraphy, which determine sleep and wakefulness by logging the timing of day-to-day touchscreen interactions [8,10–12]. While there is an ongoing debate regarding their validity, we are entering an era in which sleep assessments are becoming more accessible to users.

Observational studies investigating the relationship between nutrition and sleep often rely on subjective assessments of nutrition and/or sleep, with many highlighting the potential for recall bias [2,3]. Most analyses focus on the intake of individual nutrients; however, macronutrients that contribute to the total energy intake (such as carbohydrates, proteins, and fats, including saturated and unsaturated fatty acids) are interdependent. Although recent studies have considered these interdependencies and examined their relationship with health status [13–15], there is still insufficient research on the relationship between nutrients and sleep. In particular, these methods, which consider the interdependencies of nutrient components, are expected to attract more attention in the future than traditional methods but have not yet been fully explored [16]. Therefore, an approach that considers the relationships between these nutrients is necessary.

This cross-sectional study aimed to examine the relationships among macronutrients, dietary components, and sleep parameters. Second, we investigated the relationships between sleep and macronutrients while considering their interdependencies using compositional data analysis.

## Methods

### Study design and participants

This retrospective cross-sectional study used data from 6,052 users who consented to participate between January 19th and 31st, 2024 (Figure 1). In-app messages were sent to all “Asken” users, a dietary management and recording application, from January 19th through 31st, 2024, asking those who used both “Pokémon Sleep,” a sleep tracking application, for permission to use their stored data. Respondents were asked to provide their “Pokémon Sleep” user IDs in their electronic informed consent so that the “Pokémon Sleep” and “Asken” data could be linked. This study was approved by the Institutional Review Board of Sapporo Yurinokai Hospital, Japan.

**Figure 1.**
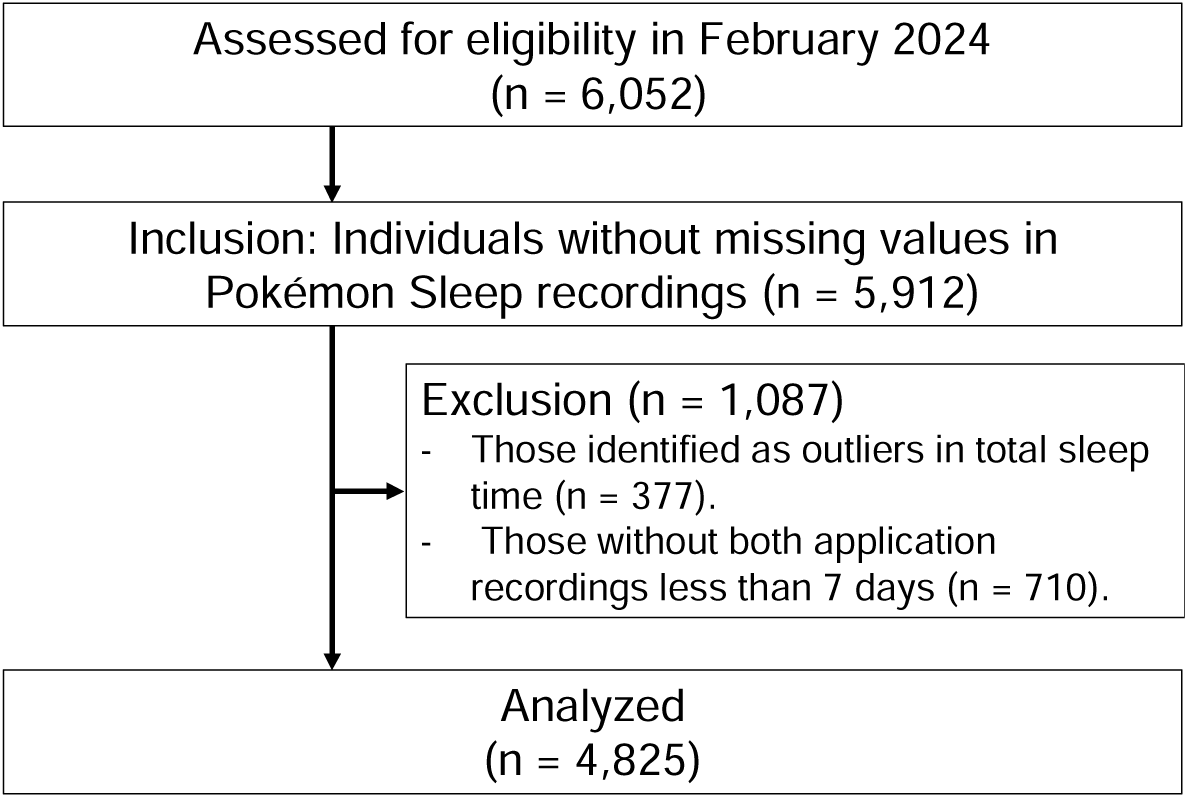
Participants flow chart

The participants used both applications concurrently from July 2023 to January 2024. Furthermore, data were extracted only from the overlapping period between the start and end dates of both applications and average values were considered representative of each participant’s typical sleep habits and nutritional intake status.

### Dietary intake patterns assessment

We calculated dietary intake patterns using the smartphone application “Asken.” The recording and calculation methods used by the participants are detailed in a previous study [9]. In this retrospective cross-sectional study, data were only included from days when participants reported consuming all three meals, as it was not possible to distinguish between missed recordings and actual skipped meals.

Asken allows for the calculation of various macronutrients and dietary components, including total energy intake, protein intake, total fat intake (including saturated, monounsaturated, and polyunsaturated fats), carbohydrate intake, sodium intake, potassium intake, calcium intake, magnesium intake, phosphorus intake, iron intake, zinc intake, copper intake, manganese intake, vitamin A intake, vitamin D intake, vitamin E intake, vitamin K intake, vitamin B1 intake, vitamin B2 intake, niacin intake, vitamin B6 intake, vitamin B12 intake, folate intake, pantothenic acid intake, vitamin C intake, cholesterol intake, dietary fiber intake, and alcohol intake.

Based on previous research [17,18], we selected and analyzed the following components reported to be associated with sleep: total energy intake, protein intake, total fat intake (including saturated, monounsaturated, and polyunsaturated fats), carbohydrate intake, sodium intake, potassium intake, and dietary fiber intake, and calculated the sodium-to-potassium ratio. Macronutrients were energy-adjusted by calculating the percentage of total energy intake. The absolute daily intake was calculated for other dietary components.

### Sleep assessment

We calculated the TST, SL, and %WASO derived from the “Pokémon Sleep,” which uses the Cole-Kripke algorithm for sleep-wake determination [19], as publicly disclosed [20]. Although accelerometer-based sleep determination integrated into smartphones lacks the ability to distinguish sleep stages [21,22], it provides a certain level of accuracy in determining sleep and wakefulness [22,23]. Although there may be a slight underestimation of SL, the overall TST and nocturnal awakenings showed good agreement with traditional actigraphy [22,23]. As previously mentioned, Natale et al. used the Cole-Kripke algorithm with smartphone accelerometers [23]. Briefly, sleep-wake determination was performed using accelerometer data with 10-s epochs per minute. The decision was based on a 7-min window of accelerometer data (from 4 min before to 2 min after the reference point (0 min)). A value greater than 1 indicates wakefulness, and a value less than 1 indicates sleep [19]. SL was defined as the time from going to bed (start of the recording) to the first instance of being determined as sleep.

### Statistical analysis

The multivariable regression analysis, isotemporal substitution model, and compositional data analysis were performed using the “lmtest,” “deltacomp,” “Compositions,” “robCompositions,” and “ggplot2” R packages, respectively [16,24–26]. All regression models were adjusted for age, sex, and BMI.

Basic characteristics, sleep status, and dietary nutrient status were described. First, multivariable regression analysis using the forced-entry method was employed to determine the association among macronutrients, dietary components, and objective sleep variables (TST, SL, and %WASO). We conducted quartile categorization for total energy intake, energy-adjusted macronutrients (protein intake, total fat intake including saturated, monounsaturated, and polyunsaturated fats, and carbohydrate intake), sodium intake, potassium intake, dietary fiber intake, and sodium-to-potassium ratio. All the quartile range information for each variable is presented in Supplementary Table S1. Dummy variables were created for each nutrient based on the first quartile.

Second, to investigate the linear relationship between macronutrient intake and sleep parameters, we employed an isotemporal substitution model and compositional data analysis [13–15]. In the isotemporal substitution model, when one macronutrient component is replaced with another on a one-to-one basis, the other components remain constant [13,14]. Compositional data analysis, in contrast, considers the overall composition and estimates the changes in sleep variables when the proportion of a specific macronutrient component is increased or decreased within the total composition [16]. Both methods are used in studies that consider the interdependencies of factors contributing to a total composition of 100% (e.g., dietary nutrient intake, 24-h physical/sedentary activity, and TST) [13,14,25,26].

We calculated the macronutrient intake (in grams) by multiplying each intake by its respective calorie contribution (i.e., 4 kcal for carbohydrates and protein and 9 kcal for each type of fat), ensuring that the sum equals 1 (i.e., total energy intake). Nutrient intake data were transformed into compositional data using an additive log-ratio transformation. The variability in the data, in terms of the variability of each macronutrient intake relative to the variability of other components and the total variance of the whole composition, is described in Supplementary Table S2 through a variation matrix within each component. A value close to zero implied that the intake of the two macronutrient components was highly proportional. The compositional data were then subjected to an isometric log-ratio (ilr) transformation to facilitate the application of the linear regression models. The ilr transformation was conducted using default and customized orthonormal bases for different nutrients. Linear regression models were constructed to evaluate the association between sleep parameters and the ilr-transformed nutrient composition after adjusting for sex, age, and BMI (Supplementary Table S3). The mean nutrient composition was calculated, and predictions for sleep parameters were made using a linear regression model. The compositional ratios of macronutrients among the participants are presented in Supplementary Table S4. Because of the isotemporal substitution model and compositional data analysis, it was not possible to replace values below the minimum composition. Therefore, in this study, we replaced values exceeding the upper limit of 6% (Supplementary Table S4).

To assess the impact of altering the nutrient composition, specific changes (e.g., increasing monounsaturated fat and decreasing polyunsaturated fat) were applied, and new predictions were obtained. Confidence intervals for the differences in the predicted sleep parameters were computed using the residual standard error, model matrix, and critical value from the t-distribution. Hypothesis tests were conducted to examine the significance of the associations between individual nutrients and sleep parameters. Different ilr transformations were applied, each focusing on a specific nutrient (protein, carbohydrate, saturated fat, monounsaturated fat, or polyunsaturated fat), and the corresponding linear models were analyzed for significance.

All descriptive and statistical analyses were performed using R 4.4.0 (R Foundation for Statistical Computing, Vienna, Austria), with statistical significance assessed at the level of 0.05.

## Results

Out of the 6,052 participants who consented, 140 individuals were excluded from the analysis due to missing sleep records, and 377 were excluded due to outliers in TST (i.e., Participants who did not fall within the average ± 3 standard deviations range.) and 710 were excluded because they used both applications for less than 7 d each. Therefore, the final analysis included 4,825 participants (mean age: 36.7 ± 10.4 years; BMI: 24.8 ± 4.8 kg/m²; 81.6% were female) (Figure 1 and Table 1).

**Table 1.** Characteristics of participants.

| Variables | All participants<br>(n =4,825) | Male<br>(n =887) | Female<br>(n = 3,938) |
| --- | --- | --- | --- |
| **Basic characteristics** |  |  |  |
| Age (years) | 36.7 ± 10.4 | 38.4 ±10.8 | 36.3 ± 10.2 |
| BMI, kg/m<sup>2</sup> | 24.8 ± 4.8 | 26.0 ± 4.5 | 24.6 ± 4.8 |
| Physical activity level, inactive, n (%) | 4,381 (90.8) | 775 (87.4) | 3,606 (91.6) |
| **Sleep status** |  |  |  |
| No. of days of Pokémon Sleep recording, d | 134.0 ± 54.5 | 135.7 ± 56.0 | 133.7 ± 54.2 |
| Total sleep time, hour | 6.7 ± 1.1 | 6.5 ± 1.1 | 6.7 ± 1.1 |
| Sleep onset latency, min | 21.5 ± 12.1 | 23.5 ± 13.6 | 21.1 ± 11.7 |
| %Wakefulness after sleep onset, % | 9.9 ± 7.2 | 13.1 ± 9.1 | 9.2 ± 6.4 |
| **Nutrients status** |  |  |  |
| No. of days of Asken data recording, d | 85.8 ± 63.6 | 93.2 ± 67.3 | 84.1 ± 62.7 |
| Total energy intake, kcal | 1662.6 ± 330.3 | 2055.1 ± 349.2 | 1574.2 ± 252.4 |
| Protein intake, % total kcal | 17.7 ± 3.7 | 18.8 ± 4.8 | 17.5 ± 3.4 |
| Carbohydrate intake, % total kcal | 51.7 ± 5.5 | 50.3 ± 5.9 | 52.0 ± 5.3 |
| Total fat intake, % total kcal | 32.3 ± 4.3 | 31.4 ± 4.8 | 32.6 ± 4.2 |
| Saturated fat intake, % total kcal | 9.5 ± 1.8 | 8.8 ± 1.8 | 9.7 ± 1.8 |
| Monounsaturated fat intake, % total kcal | 11.5 ± 2.0 | 11.3 ± 2.2 | 11.5 ± 1.9 |
| Polyunsaturated fat intake, % total kcal | 6.3 ± 1.0 | 6.1 ± 1.2 | 6.3 ± 1.0 |
| Dietary fiber intake, g/d | 11.7 ± 3.5 | 10.7 ± 2.7 | 11.9 ± 3.6 |
| Sodium, mg/d | 3486.6 ± 881.0 | 4230.7 ± 996.7 | 3319.1 ± 758.1 |
| Potassium, mg/d | 2295.3 ± 602.1 | 2625.0 ± 692.4 | 2221.0 ± 553.3 |
| Sodium to Potassium ratio, ratio | 1.6 ± 0.4 | 1.7 ± 0.5 | 1.6 ± 0.4 |

Note: BMI, body mass index

The macronutrient composition was as follows: protein, 18.2%; carbohydrates, 53.8%; and total fat, 28.0% (saturated fat at 9.7%, monounsaturated fat at 11.8%, and polyunsaturated fat at 6.5%) (Supplementary Table S4).

Figure 2 and Supplementary Table S5 present the results of multivariate regression analyses comparing each nutrient across quartiles to the first quartile in relation to sleep variables. In terms of macronutrients, TST was significantly shorter in the second, third, and fourth quartiles than that in the group with lower total energy intake. Higher intake ratios of total fat, including saturated, monounsaturated, and polyunsaturated fats, were associated with shorter TST. However, higher protein intake levels in the third and fourth quartiles were associated with longer TST than those in the first quartile. Higher intake levels of total fat, saturated fat, and monounsaturated fat were associated with longer SL. However, a higher intake of polyunsaturated fats showed a tendency towards shorter SL. Higher total energy and total fat intakes (including saturated and monounsaturated fats) were associated with longer %WASO. Conversely, a higher carbohydrate intake showed a tendency towards a shorter %WASO.

**Figure 2.**
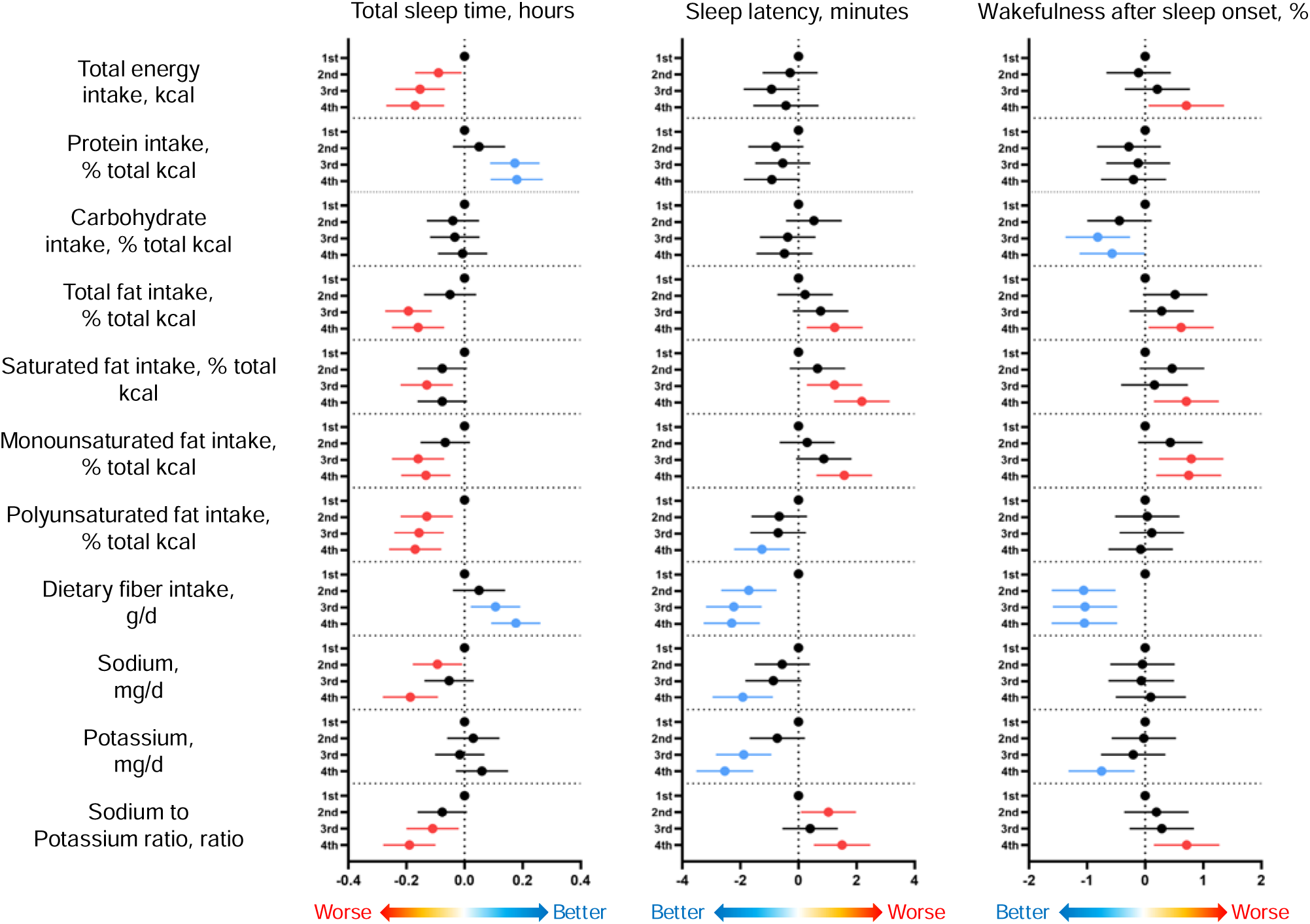
Multivariable regression analysis of macronutrients and dietary components on sleep variables

Higher dietary fiber intake was consistently associated with longer TST, shorter SL, and %WASO, indicating that fiber is a beneficial nutrient for sleep. Higher sodium intake was associated with shorter TST. Higher sodium and potassium intake was associated with shorter SL. Additionally, higher potassium intake was associated with a shorter %WASO. However, a higher sodium-to-potassium ratio was associated with shorter TST, longer SL, and %WASO (Figure 2, Supplementary Table S5).

The results of the isotemporal substitution model showed that replacing 6% of total energy intake from protein with carbohydrates was significantly associated with a reduction of 0.2 h in TST. Conversely, replacing 6% of polyunsaturated fat with equivalent amounts of protein, carbohydrates, saturated fat, and monounsaturated fat was significantly associated with increases in TST by 0.6, 0.4, 0.5, and 0.6 h, respectively (Figure 3 and Supplementary Table S6). Regarding SL, replacing 6% of proteins or carbohydrates with monounsaturated fat was significantly associated with an increase of 2.4 and 2.8 min, respectively. Conversely, replacing 6% of monounsaturated fat with equivalent amounts of protein, carbohydrates, and saturated fat was significantly associated with SL reductions of 3.7, 4.1, and 3.2 min, respectively. Additionally, replacing 6% polyunsaturated fat with equivalent amounts of protein, carbohydrates, saturated fat, and monounsaturated fat was significantly associated with an increase in SL of 10.3, 9.9, 10.8, and 12.7 min, respectively (Figure 4 and Supplementary Table S6). For %WASO, replacing 6% of protein with monounsaturated fat was significantly associated with a 1.2% increase, whereas replacing saturated fat with monounsaturated fat was significantly associated with a 2.3% increase. Conversely, replacing 6% of monounsaturated fat with equivalent amounts of protein, carbohydrates, and saturated fat was significantly associated with a reduction in %WASO by 1.8%, 1.7%, and 2.5%, respectively. Additionally, replacing 6% polyunsaturated fat with equivalent amounts of protein, carbohydrates, saturated fat, and monounsaturated fat was significantly associated with an increase in %WASO of 4.3%, 4.5%, 3.7%, and 5.5%, respectively (Figure 5 and Supplementary Table S6).

**Figure 3.**
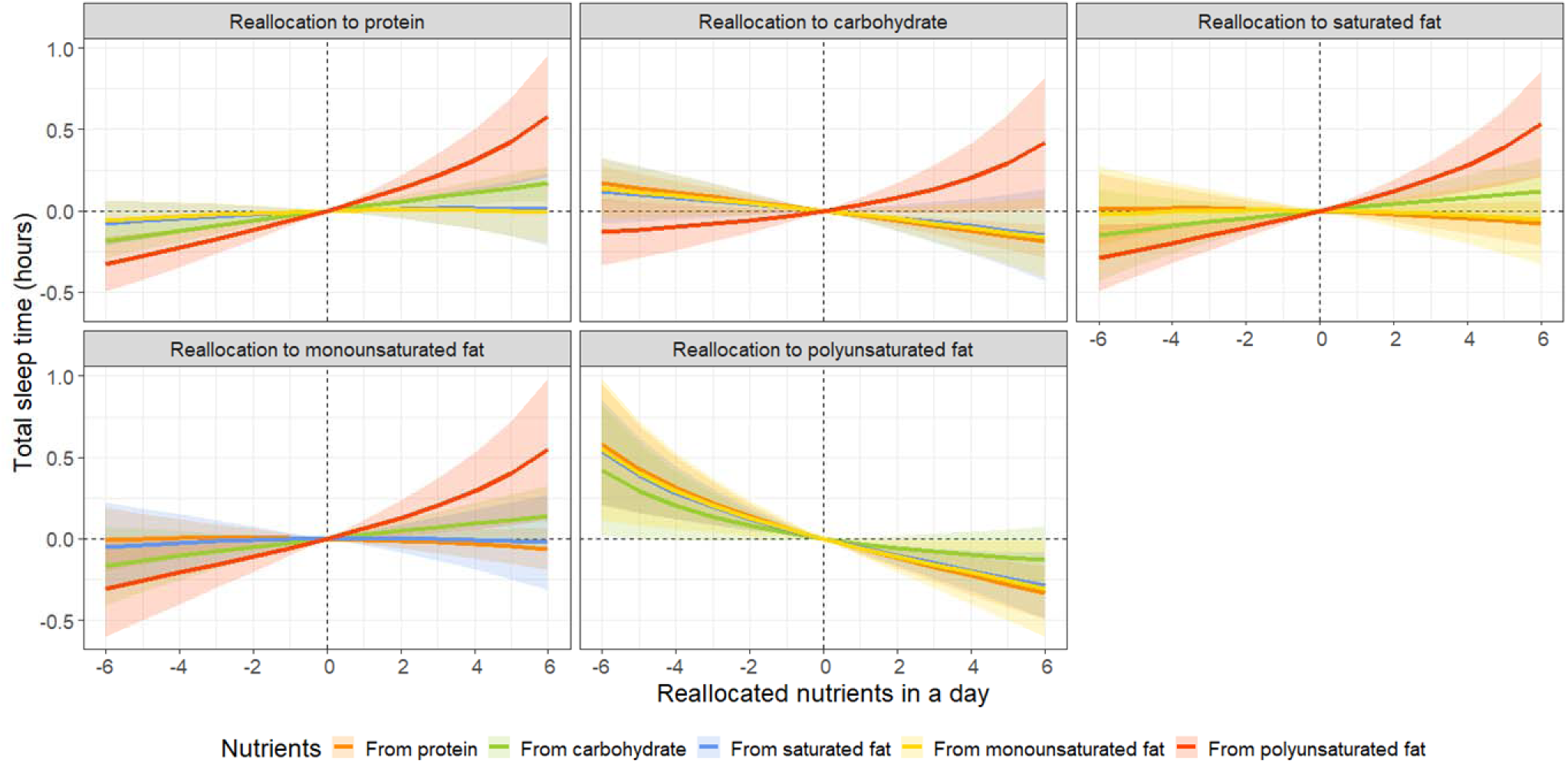
Simulated changes in total sleep time when reallocating fixed amounts of other nutrient components among each nutrient component, while keeping the remaining components constant at compositional percentages. All estimates have been adjusted for age, sex, body mass index.

**Figure 4.**
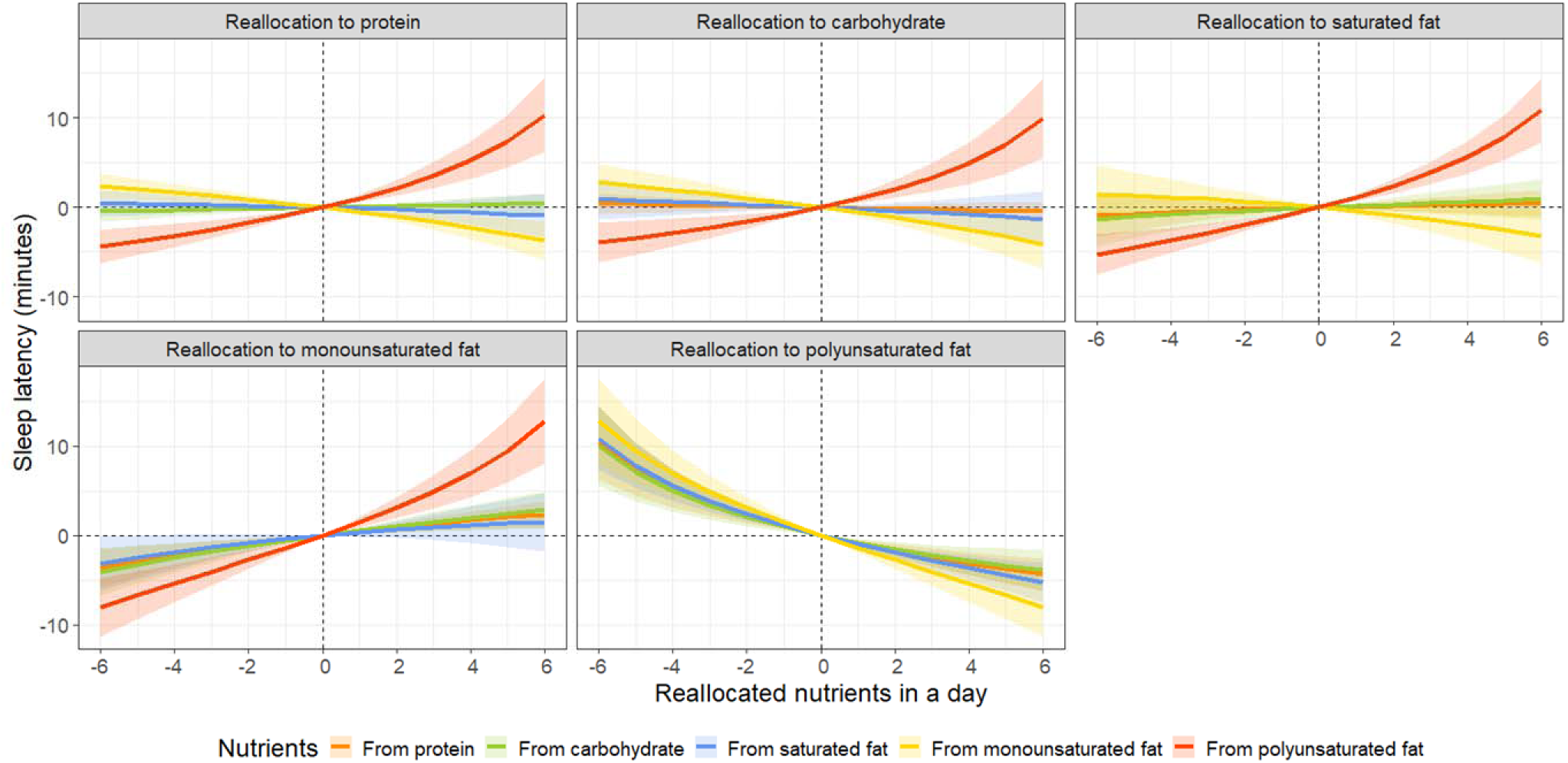
Simulated changes in sleep latency when reallocating fixed amounts of other nutrient components among each nutrient component, while keeping the remaining components constant at compositional percentages. All estimates have been adjusted for age, sex, body mass index.

**Figure 5.**
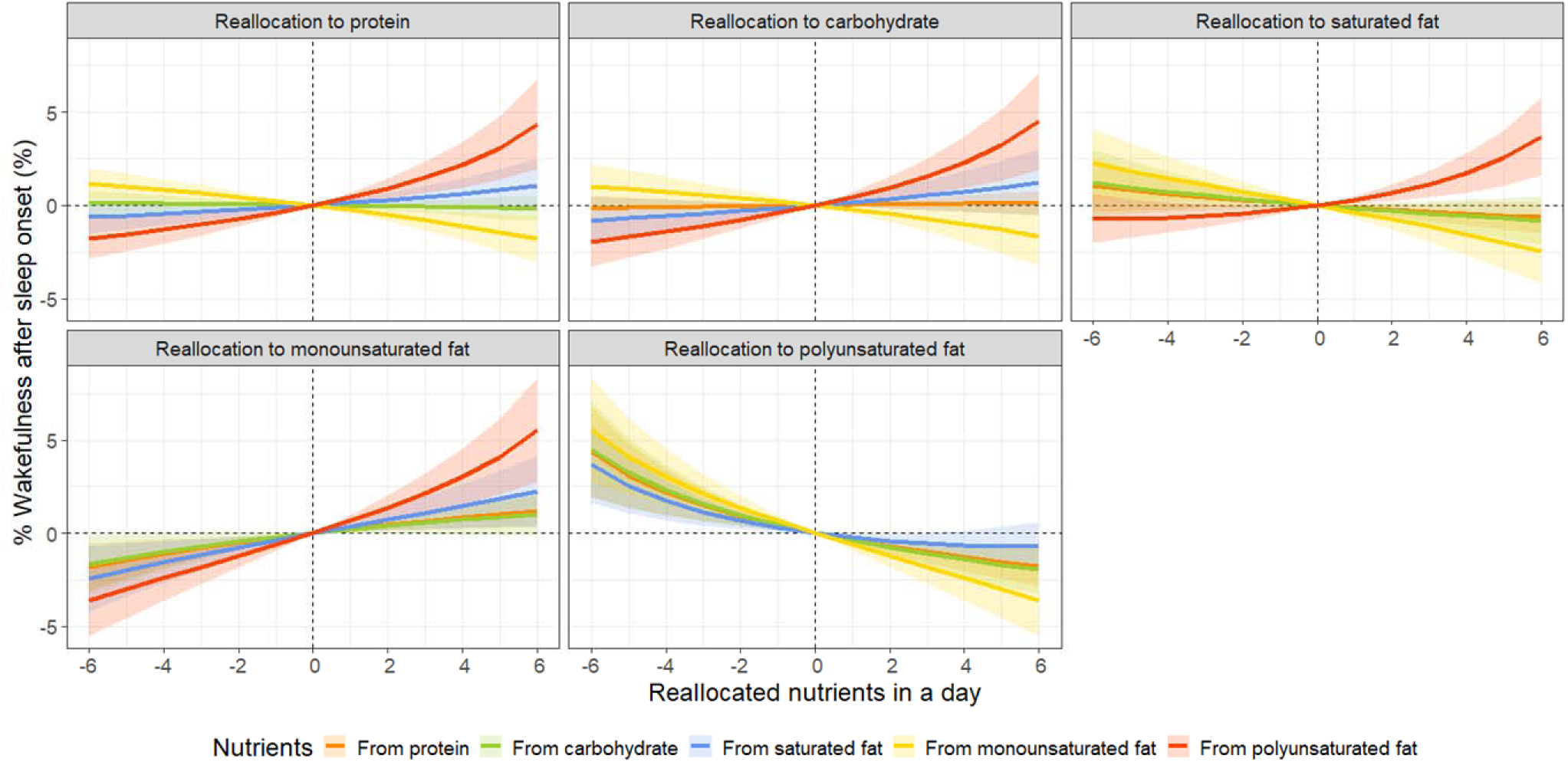
Simulated changes in the proportion of wakefulness after sleep onset when reallocating fixed amounts of other nutrient components among each nutrient component, while keeping the remaining components constant at compositional percentages. All estimates have been adjusted for age, sex, body mass index.

In the compositional data analysis, reducing protein by 6% of the overall macronutrient composition was significantly associated with a decrease in the TST by 0.6 h, whereas reducing polyunsaturated fat by 6% was significantly associated with an increase in the TST by 0.5 h. Conversely, increasing protein by 6% from the overall macronutrient composition was significantly associated with an increase in TST by 0.3 h, while increasing polyunsaturated fat by 6% was significantly associated with a decrease in TST by 0.2 h (Table 2). Regarding SL, reducing monounsaturated fat by 6% from the overall macronutrient composition was significantly associated with a decrease of 10.5 min, whereas reducing polyunsaturated fat by 6% was associated with an increase of 10.7 min. Conversely, increasing monounsaturated fat by 6% was significantly associated with an increase of 4.6 min, and increasing polyunsaturated fat by 6% was significantly associated with a decrease of 4.7 min (Table 2). For %WASO, similar to SL, reducing monounsaturated fats by 6% of the overall macronutrient composition was significantly associated with a decrease of 5.0%, whereas a 6% reduction in polyunsaturated fats was significantly associated with an increase of 4.5%. Conversely, increasing the monounsaturated fat content by 6% was significantly associated with an increase of 2.2%, and increasing the polyunsaturated fat content by 6% was significantly associated with a decrease of 2.0% (Table 2).

**Table 2.** Estimated non-standardized coefficients for displacing 6% of macronutrients between one and the remaining macronutrients proportionally.

|  | Total sleep time, hours |  | Sleep latency, minutes |  | % Wakefulness after sleep onset, % |  |
| --- | --- | --- | --- | --- | --- | --- |
|  | Take 6% from one nutrient and distribute it. | Add 6% to one nutrient from the others. | Take 6% from one nutrient and distribute it. | Add 6% to one nutrient from the others. | Take 6% from one nutrient and distribute it. | Add 6% to one nutrient from the others. |
| Protein | **-0.61**<br>**(-0.80, -0.41)** | **+0.27**<br>**(0.18, 0.35)** | -0.78<br>(-2.93, 1.38) | +0.34<br>(-0.61, 1.29) | -0.06<br>(-1.32, 1.20) | +0.03<br>(-0.53, 0.58) |
| Carbohydrate | +0.60<br>(-0.20, 1.40) | -0.27<br>(-0.62, 0.09) | +2.79<br>(-5.98, 11.57) | -1.23<br>(-5.10, 2.64) | -1.70<br>(-6.81, 3.41) | +0.75<br>(-1.51, 3.01) |
| Saturated fat | -0.20<br>(-0.67, 0.28) | +0.09<br>(-0.12, 0.30) | -2.20<br>(-7.40, 2.99) | +0.97<br>(-1.32, 3.26) | +2.23 (-0.80, 5.26) | -0.99<br>(-2.32, 0.35) |
| Monounsaturated fat | -0.30<br>(-0.85, 0.26) | +0.13<br>(-0.11, 0.38) | **-10.50**<br>**(-16.63, -4.38)** | **+4.64**<br>**(1.93, 7.34)** | **-5.00**<br>**(-8.57, -1.43)** | **+2.21**<br>**(0.63, 3.78)** |
| Polyunsaturated fat | **+0.50**<br>**(0.12, 0.88)** | **-0.22**<br>**(-0.39, -0.05)** | **+10.69**<br>**(6.48, 14.90)** | **-4.72**<br>**(-6.58, -2.86)** | **+4.53**<br>**(2.07, 6.99)** | **-2.00**<br>**(-3.08, -0.92)** |

Note: Data presented as non-standardized coefficients (95% confidence interval). All estimates have been adjusted for age, sex, body mass index. The results in bold are significant ( $P < .05$ ). Non-standardized coefficients reflect the estimated change in sleep parameters associated with reallocating 6% of macronutrients between one and the remaining macronutrients proportionally.

## Discussion

This study examined the relationship between nutritional intake and sleep using multivariate regression analysis, isotemporal substitution models, and compositional data analysis. Overall, the findings are consistent with previous research [3,4]. Higher total energy, fat, and sodium intakes were associated with shorter TST, while higher protein and dietary fiber intakes were associated with longer TST. Dietary fiber intake was consistently associated with improved SL and %WASO. Additionally, higher sodium intake was correlated with shorter TST, whereas higher potassium intake was associated with shorter SL. Notably, polyunsaturated fats demonstrated potential benefits for SL and %WASO compared to other types of fat.

Studies suggest that dietary fiber can affect sleep by influencing microbiota [27]. When dietary fibers are fermented by microbiota in the large intestine, they produce short-chain fatty acids, such as acetate, propionate, and butyrate, which enhance serotonin release. Furthermore, administering tributyrin, a compound made of three molecules of butyric acid and glycerol, to mice resulted in a 50% increase in non-rapid eye movement sleep for 4 h [28]. The results of this study support those of previous studies [27,28].

Previous studies have established that higher total energy intake and poor dietary quality are associated with lower subjective sleep quality [5,29]. Specifically, higher protein intake correlates with improved sleep, whereas a higher intake of fatty acids is linked to worse sleep [29]. This study also found variations in the effects of lipid intake on sleep, depending on the type (e.g., saturated, monounsaturated, or polyunsaturated fats). The mechanism by which proteins affect sleep following acute feeding may involve tryptophan, tyrosine, and the synthesis of brain neurotransmitters, such as serotonin, melatonin, and dopamine [30]. Although no significant differences were observed in SL or %WASO, a higher protein intake was positively associated with longer TST.

However, the specific mechanisms underlying the relationship among monounsaturated fat, polyunsaturated fat, and sleep remain unclear. A meta-analysis found an association between a higher intake of hexadecenoic acid, a saturated fatty acid, and difficulty falling asleep [31]. Conversely, difficulties falling asleep were linked to a lower intake of dodecanoic acid, a monounsaturated fat, and butanoic acid, a saturated fatty acid. Additionally, a reduced intake of dodecanoic acid was associated with difficulties in maintaining sleep. An epidemiological study involving adults evaluated the relationship between two polyunsaturated fatty acids, omega-6 and omega-3, and found that omega-6 fatty acids were related to sleep disorder risk in an inverse U-shaped manner [32]. The omega-6/omega-3 ratio was positively and linearly associated with the risk of sleep disorders. The underlying mechanisms involving omega-3 fatty acids and sleep are discussed, noting that a deficiency in omega-3 intake may affect cortical neuron oscillatory activity and sleep-wake patterns [33].

Additionally, the timing of fat intake may influence sleep; one observational study based on polysomnography suggested that consuming high-fat foods close to bedtime may be associated with increased WASO [34]. Continued consumption of a high-fat diet has been partially understood to reduce physical activity and increase rapid eye movements and sleep fragmentation through dopaminergic dysregulation [35]. Research involving mouse models and pregnant women have suggested a potential positive association between monounsaturated fat intake and sleep [36,37]. However, other studies have indicated a negative association between monounsaturated fat and sleep, whereas polyunsaturated fat has shown a positive association with sleep [38–40]. Previous studies have often focused on dietary fiber levels and comparisons between high- and low-saturated-fat diets [35,38]. Future studies should explore the impact of unsaturated fats, including monounsaturated and polyunsaturated fats.

Sodium and potassium have been reported to impact health individually, and a high sodium-to-potassium ratio (indicating relatively high sodium intake) has been associated with higher all-cause mortality [41]. Sodium intake is known to be correlated with sleep, indicating that higher sodium intake is associated with shorter TST and worse sleep quality [18,40]. This relationship is attributable to the potential effect of sodium on nocturnal urination [42,43]. However, a higher intake of sodium alone was associated with a shorter total sleep time, although no significant differences were observed in %WASO. Moreover, greater sodium intake was associated with shorter SL. Higher potassium intake was associated with shorter SL and reduced %WASO. All sleep variables were adversely related when considering the balance between sodium and potassium, which indicates a healthy dietary pattern (i.e., relatively higher sodium intake). In this study’s multivariable regression analysis, the grouping was based on absolute sodium intake without considering the relative amount of potassium. Therefore, the results linking sleep outcomes with the sodium-to-potassium ratio are considered reasonable, as they provide a more comprehensive perspective on the impact of both minerals on sleep.

In this study, we investigated the association between daily nutrient intake and sleep using isotemporal substitution models and compositional data analyses to account for the interdependencies [16]. These approaches enabled us to assess how substituting macronutrients and considering the overall composition ratio affect sleep outcomes. Nonetheless, this study had several limitations. First, this cross-sectional study does not provide causal explanations. As noted above, previous studies have reported bidirectional relationships between these factors. Additionally, the isotemporal substitution model and compositional data analysis are merely statistical inferences [16]. The findings of this study underscore the need for future interventions and longitudinal studies to establish causal effects definitively. Second, this study may have included a highly health-conscious group that uses both health management applications simultaneously. Therefore, although values around the 2nd and 3rd quartiles, as shown in Supplementary Table 2, correspond to Japanese macronutrient standards [44] and suggest the possibility of representing the general population, caution is needed when generalizing the findings to broader populations, owing to the inclusion of this health-conscious group. Third, the sleep assessment method used in this study involved applying gamified elements. Some users may have exhibited sleep patterns different from their habitual patterns to achieve specific goals within the game. Moreover, because users have their own smartphones, the reliability of accelerometer measurements varies across different device models. In the future, integrating alternative methods, such as tappigraphy, for sleep-wake rhythms, actigraphy, or portable EEG devices could provide more objective evaluations [10–12]. Finally, because this study was retrospective, we could not adequately capture confounding factors related to sleep and nutrition. We collected basic demographic information, such as age, sex, height, weight, and physical activity level. However, other confounding factors, such as alcohol and smoking habits, employment status (including shifts and night shifts), marital status, cohabitation status, medical history, and medication use, were not considered.

## Conclusions

We found that a higher intake of polyunsaturated fats was associated with better SL and wakefulness, whereas saturated and monounsaturated fats were correlated with longer SL and wakefulness. Dietary fiber intake was positively associated with sleep quality, whereas a higher sodium-to-potassium ratio was associated with worse sleep. Protein intake was associated with longer TST, and monounsaturated fat intake was associated with longer SL and wakefulness. Despite its cross-sectional nature, this study highlights the intricate role of dietary factors in sleep, suggesting the potential for dietary interventions to enhance sleep health.

## Data Availability

The data used in this study belong to The Pokemon Company (Tokyo, Japan) and Asken Inc. (Tokyo, Japan). These data are not publicly available but can be used in future projects. Proposals and requests for data access were sent to the corresponding authors via email.

## Acknowledgments

We express our gratitude to the personnel of The Pokémon Company (Tokyo, Japan), Asken Inc. (Tokyo, Japan), and S’UIMIN Inc. (Tokyo, Japan) for their contributions to data preparation. We also thank Editage (www.editage.com) for the English language editing.

## Author Contributions

J. S., M. I., and Y. M.; methodology, J. S., M. I., and M. K.; formal analysis, J. S. and M. I.; writing—original draft preparation, J. S.; writing—review and editing, J. S., M. I., M. K., and Y. M.; visualization, J. S.; supervision, M. I. and Y. M.; funding acquisition, Y. M. All authors have read and agreed to the published version of the manuscript.

## Institutional Review Board Statement

This study was conducted in accordance with the Declaration of Helsinki and approved by the Institutional Review Board of Sapporo Yuurinokai Hospital, Japan.

## Informed Consent Statement

Informed consent was obtained from all participants involved in the study.

## Data Availability Statement

The data used in this study belong to The Pokémon Company (Tokyo, Japan) and Asken Inc. (Tokyo, Japan). These data are not publicly available but can be used in future projects. Proposals and requests for data access were sent to the corresponding authors via email.

## Abbreviations

95% CI: 95% confidence interval
BMI: body mass index
ilr: isometric log-ratio
%WASO: % wakefulness after sleep onset
SL: sleep latency
TST: total sleep time

