## Supplementary Table S1-S6 for "Relationship between macronutrients, dietary components, and objective sleep variables measured by smartphone applications"

Correspondence should be addressed to Masashi Yanagisawa

This PDF file includes:

Table S1 and S6

**Table S1**. Range of quartiles for macronutrient and dietary components in multivariable regression analysis

|  | 1^st^ quartile  (n = 1,207) | 2^nd^ quartile  (n = 1,206) | 3^rd^ quartile  (n = 1,206) | 4^th^ quartile  (n = 1,206) |
| --- | --- | --- | --- | --- |
| Total energy intake,  kcal | 369.13–1450.56 | 1450.66–1619.27 | 1619.32–1834.00 | 1834.04–5019.60 |
| Protein intake,  % total kcal | 8.31–15.24 | 15.25–16.86 | 16.87–19.27 | 19.28–48.37 |
| Carbohydrate intake,  % total kcal | 14.14–48.74 | 48.75–52.08 | 52.08–55.12 | 55.12–84.84 |
| Total fat intake,  % total kcal | 15.29–29.68 | 29.69–32.43 | 32.44–35.01 | 35.02–61.33 |
| Saturated fat intake,  % total kcal | 2.34–8.31 | 8.32–9.48 | 9.48–10.67 | 10.67–19.29 |
| Monounsaturated fat intake,  % total kcal | 2.65–10.28 | 10.29–11.54 | 11.54–12.74 | 12.74–24.64 |
| Polyunsaturated fat intake,  % total kcal | 1.34–5.63 | 5.63–6.25 | 6.26–6.89 | 6.89–13.61 |
| Dietary fiber intake,  g/d | 2.65–9.61 | 9.62–11.04 | 11.05–12.85 | 12.85–104.21 |
| Sodium,  mg/d | 406.80–2918.86 | 2919.01–3405.71 | 3406.36–3959.13 | 3959.18–11506.16 |
| Potassium,  mg/d | 333.31–1889.13 | 1889.45–2232.65 | 2232.71–2635.82 | 2636.15–6133.66 |
| Sodium to Potassium ratio, ratio | 0.19–1.30 | 1.30–1.54 | 1.55–1.81 | 1.81–4.87 |

**Table S2.** Compositional variation matrix of macronutrient components

| Variables | Protein | Carbohydrate | Saturated fat | Monounsaturated fat | Polyunsaturated fat |
| --- | --- | --- | --- | --- | --- |
| Protein | 0 | 0.064 | 0.101 | 0.085 | 0.064 |
| Carbohydrate | 0.064 | 0 | 0.013 | 0.008 | 0.019 |
| Saturated fat | 0.101 | 0.013 | 0 | 0.016 | 0.047 |
| Monounsaturated fat | 0.085 | 0.008 | 0.016 | 0 | 0.023 |
| Polyunsaturated fat | 0.064 | 0.019 | 0.047 | 0.023 | 0 |

Note: A value close to zero implies that the intake of the two macronutrient components involved in the ratio is highly proportional.

**Table S3.** Association of macronutrient components with sleep parameters in study participants

|  | Total sleep time, hours | | Sleep latency, minutes | | % Wakefulness after sleep onset (%) | |
| --- | --- | --- | --- | --- | --- | --- |
| Variables | B | (95%CI) | B | (95%CI) | B | (95%CI) |
| Protein,  % total kcal | **0.48** | **(0.33, 0.64)** | 0.62 | (-1.10, 2.33) | 0.05 | (-0.95, 1.05) |
| Carbohydrate,  % total kcal | -0.48 | (-1.11, 0.16) | -2.22 | (-9.19, 4.75) | -1.48 | (-2.71, 5.41) |
| Saturated fat,  % total kcal | 0.16 | (-0.22, 0.53) | 1.75 | (-2.38, 5.88) | -1.77 | (-4.18, 0.63) |
| Monounsaturated fat,  % total kcal | 0.24 | (-0.21, 0.68) | **8.35** | **(3.48, 13.21)** | **3.97** | **(1.14, 6.81)** |
| Polyunsaturated fat,  % total kcal | **-0.40** | **(-0.70, -0.09)** | **-8.49** | **(-11.84, -5.15)** | **-3.60** | **(-5.55, -1.65)** |

Note: Data are presented as non-standardized coefficients with 95% confidence intervals. The results in bold are significant (*P* < .05). All estimates have been adjusted for age, sex, body mass index.

|  | Mean | Min | Max |
| --- | --- | --- | --- |
| Protein, % total kcal | 18.2 | 8.5 | 45.4 |
| Carbohydrate, % total kcal | 53.8 | 15.1 | 76.9 |
| Saturated fat, % total kcal | 9.7 | 2.8 | 20.5 |
| Monounsaturated fat, % total kcal | 11.8 | 3.1 | 26.2 |
| Polyunsaturated fat, % total kcal | 6.5 | 1.7 | 13.8 |

**Table S4**. Compositional ratios of macronutrients among participants

|  |  | Total sleep time, hour | | Sleep latency, min | | % Wakefulness after sleep onset, % | |
| --- | --- | --- | --- | --- | --- | --- | --- |
|  |  | Unadjusted | Sex, age, and BMI adjusted | Unadjusted | Sex, age, and BMI adjusted | Unadjusted | Sex, age, and BMI adjusted |
| Total energy intake, kcal | 2^nd^ quartile | **-0.09 (-0.18, -0.01)** | **-0.09 (-0.17, -0.01)** | -0.19 (-1.15, 0.78) | -0.29 (-1.24, 0.66) | -0.01 (-0.57, 0.56) | -0.11 (-0.67, 0.44) |
|  | 3^rd^ quartile | **-0.18 (-0.26, -0.09)** | **-0.15 (-0.24, -0.07)** | -0.38 (-1.34, 0.59) | -0.93 (-1.89, 0.03) | **0.73 (0.17, 1.29)** | 0.21 (-0.35, 0.77) |
|  | 4^th^ quartile | **-0.23 (-0.32, -0.15)** | **-0.17 (-0.27, -0.07)** | **1.55 (0.58, 2.51)** | -0.43 (-1.56, 0.69) | **2.88 (2.32, 3.44)** | **0.71 (0.06, 1.36)** |
| Protein intake, % total kcal | 2^nd^ quartile | 0.04 (-0.05, 0.13) | 0.05 (-0.04, 0.14) | -0.69 (-1.65, 0.28) | -0.78 (-1.73, 0.17) | -0.20 (-0.77, 0.38) | -0.28 (-0.83, 0.27) |
|  | 3^rd^ quartile | **0.17 (0.08, 0.26)** | **0.17 (0.09, 0.26)** | -0.38 (-1.35, 0.58) | -0.54 (-1.49, 0.41) | -0.05 (-0.62, 0.52) | -0.12 (-0.67, 0.43) |
|  | 4^th^ quartile | **0.17 (0.08, 0.25)** | **0.18 (0.09, 0.27)** | -0.09 (-1.06, 0.87) | -0.92 (-1.88, 0.05) | 0.34 (-0.24, 0.91) | -0.20 (-0.76, 0.36) |
| Carbohydrate　intake, % total kcal | 2^nd^ quartile | -0.01 (-0.10, 0.08) | -0.04 (-0.13, 0.05) | 0.27 (-0.70, 1.24) | 0.53 (-0.43, 1.49) | **-0.89 (-1.46, -0.32)** | -0.44 (-1.00, 0.11) |
|  | 3^rd^ quartile | 0.01 (-0.08, 0.09) | -0.03 (-0.12, 0.05) | -0.75 (-1.71, 0.22) | -0.37 (-1.33, 0.59) | **-1.35 (-1.92, -0.78)** | **-0.82 (-1.37, -0.26)** |
|  | 4^th^ quartile | 0.05 (-0.04, 0.13) | -0.01 (-0.09, 0.08) | **-1.21 (-2.18, -0.25)** | -0.49 (-1.45, 0.48) | **-1.41 (-1.98, -0.84)** | **-0.57 (-1.13, -0.01)** |
| Total fat intake,  % total kcal | 2^nd^ quartile | -0.06 (-0.15, 0.03) | -0.05 (-0.14, 0.04) | 0.03 (-0.65, 1.28) | 0.23 (-0.73, 1.18) | 0.54 (-0.03, 1.11) | 0.52 (-0.04, 1.07) |
|  | 3^rd^ quartile | **-0.11 (-0.20, -0.24)** | **-0.11 (-0.20, -0.27)** | 0.93 (-0.04, 1.89) | 0.76 (-0.19, 1.72) | 0.17 (-0.40, 0.74) | 0.28 (-0.27, 0.84) |
|  | 4^th^ quartile | **-0.16 (-0.25, -0.08)** | **-0.16 (-0.25, -0.07)** | **1.60 (0.63, 2.56)** | **1.25 (0.28, 2.21)** | **0.62 (0.05, 1.19)** | **0.62 (0.06, 1.18)** |
| Saturated fat intake, % total kcal | 2^nd^ quartile | -0.07 (-0.16, 0.02) | -0.08 (-0.16, 0.01) | 0.54 (-0.43, 1.50) | 0.65 (-0.30, 1.61) | 0.23 (-0.34, 0.80) | 0.46 (-0.09, 1.02) |
|  | 3^rd^ quartile | **-0.11 (-0.20, -0.02)** | **-0.13 (-0.22, -0.04)** | **1.16 (0.19, 2.12)** | **1.24 (0.29, 2.20)** | -0.25 (-0.82, 0.32) | 0.15 (-0.41, 0.74) |
|  | 4^th^ quartile | -0.05 (-0.14, 0.04) | -0.08 (-0.16, 0.01) | **2.04 (1.08, 3.01)** | **2.18 (1.22, 3.14)** | 0.19 (-0.38, 0.77) | **0.71 (0.15, 1.27)** |
| Monounsaturated fat intake,  % total kcal | 2^nd^ quartile | -0.07 (-0.16, 0.02) | -0.07 (-0.15, 0.02) | 0.48 (-0.49, 1.44) | 0.30 (-0.65, 1.25) | 0.48 (-0.09, 1.05) | 0.43 (-0.12, 0.99) |
|  | 3^rd^ quartile | **-0.16 (-0.25, -0.08)** | **-0.16 (-0.25, -0.07)** | **1.13 (0.17, 2.10)** | 0.87 (-0.08, 1.83) | **0.84 (0.27, 1.41)** | **0.79 (0.24, 1.35)** |
|  | 4^th^ quartile | **-0.14 (-0.23, -0.06)** | **-0.13 (-0.22, -0.05)** | **2.22 (1.25, 3.18)** | **1.58 (0.62, 2.54)** | **0.96 (0.39, 1.53)** | **0.75 (0.19, 1.31)** |
| Polyunsaturated fat intake,  % total kcal | 2^nd^ quartile | **-0.13 (-0.22, -0.05)** | **-0.13 (-0.22, -0.04)** | -0.68 (-1.65, 0.29) | -0.66 (-1.62, 0.29) | -0.05 (-0.62, 0.52) | 0.04 (-0.52, 0.59) |
|  | 3^rd^ quartile | **-0.16 (-0.25, -0.07)** | **-0.16 (-0.24, -0.07)** | -0.62 (-1.59, 0.35) | -0.70 (-1.66, 0.26) | 0.04 (-0.53, 0.61) | 0.11 (-0.44, 0.67) |
|  | 4^th^ quartile | **-0.18 (-0.27, -0.09)** | **-0.17 (-0.26, -0.08)** | **-1.22 (-2.19, -0.25)** | **-1.26 (-2.22, -0.30)** | -0.13 (-0.70, 0.44) | -0.08 (-0.63, 0.48) |
| Dietary fiber intake, g/d | 2^nd^ quartile | 0.06 (-0.03, 0.15) | 0.05 (-0.04, 0.14) | **-1.99 (-2.96, -1.03)** | **-1.71 (-2.66, -0.76)** | **-1.38 (-1.94, -0.81)** | **-1.06 (-1.61, -0.51)** |
|  | 3^rd^ quartile | **0.12 (0.03, 0.21)** | **0.11 (0.02, 0.19)** | **-2.88 (-3.85, -1.92)** | **-2.23 (-3.19, -1.27)** | **-1.65 (-2.21, -1.08)** | **-1.04 (-1.59, -0.48)** |
|  | 4^th^ quartile | **0.19 (0.11, 0.28)** | **0.18 (0.09, 0.26)** | **-3.21 (-4.17, -2.25)** | **-2.30 (-3.27, -1.33)** | **-1.84 (-2.41, -1.27)** | **-1.05 (-1.61, -0.48)** |
| Sodium, mg/d | 2^nd^ quartile | **-0.10 (-0.19, -0.02)** | **-0.09 (-0.18, -0.01)** | -0.10 (-1.07, 0.86) | -0.56 (-1.51, 0.39) | 0.27 (-0.29, 0.84) | -0.05 (-0.60, 0.51) |
|  | 3^rd^ quartile | -0.08 (-0.17, 0.01) | -0.05 (-0.14, 0.03) | 0.01 (-0.96, 0.98) | -0.87 (-1.83, 0.10) | **0.68 (0.11, 1.24)** | -0.07 (-0.63, 0.50) |
|  | 4^th^ quartile | **-0.25 (-0.34, -0.16)** | **-0.19 (-0.28, -0.09)** | 0.10 (-0.87, 1.07) | **-1.92 (-2.96, -0.88)** | **2.06 (1.50, 2.63)** | 0.10 (-0.51, 0.70) |
| Potassium, mg/d | 2^nd^ quartile | 0.02 (-0.07, 0.10) | 0.03 (-0.06, 0.12) | -0.52 (-1.48, 0.45) | -0.73 (-1.68, 0.22) | 0.23 (-0.35, 0.80) | -0.02 (-0.58, 0.53) |
|  | 3^rd^ quartile | -0.05 (-0.13, 0.04) | -0.02 (-0.10, 0.07) | **-1.61 (-2.57, -0.64)** | **-1.89 (-2.84, -0.93)** | 0.27 (-0.30, 0.84) | -0.21 (-0.76, 0.35) |
|  | 4^th^ quartile | -0.01 (-0.09, 0.08) | 0.06 (-0.03, 0.15) | **-1.82 (-2.78, -0.85)** | **-2.54 (-3.52, -1.56)** | 0.42 (-0.15, 1.00) | **-0.75 (-1.32, -0.18)** |
| Sodium to Potassium ratio, ratio | 2^nd^ quartile | **-0.09 (-0.18, -0.01)** | -0.08 (-0.16, 0.01) | **1.30 (0.33, 2.26)** | **1.03 (0.08, 1.98)** | 0.41 (-0.16, 0.98) | 0.19 (-0.36, 0.75) |
|  | 3^rd^ quartile | **-0.12 (-0.21, -0.04)** | **-0.11 (-0.20, -0.02)** | **0.99 (0.03, 1.95)** | 0.40 (-0.55, 1.36) | **0.75 (0.18, 1.32)** | 0.29 (-0.27, 0.84) |
|  | 4^th^ quartile | **-0.21 (-0.29, -0.12)** | **-0.19 (-0.28, -0.10)** | **2.53 (1.57, 3.50)** | **1.50 (0.53, 2.47)** | **1.45 (0.88, 2.02)** | **0.71 (0.15, 1.28)** |

**Table S5**. Multivariable regression analysis of macronutrients and dietary components on sleep variables

Note: Data are presented as non-standardized coefficients with 95% confidence intervals. The results in bold are significant (*P* < .05).

**Table S6**. Changes in sleep parameters when reallocating fixed amounts of other nutrient components among each nutrient component, while keeping the remaining components constant at compositional percentages.

| ±6% | To protein | To carbohydrate | To saturated fat | To monounsaturated fat | To polyunsaturated fat |
| --- | --- | --- | --- | --- | --- |
| **Total sleep time, hours** | | | | | |
| From protein | Drop | **-0.19**  **(-0.29, -0.09)** | -0.08  (-0.21, 0.05) | -0.06  (-0.19, 0.07) | **-0.33**  **(-0.50, -0.17)** |
| From carbohydrate | **+0.17**  **(0.06, 0.27)** | Drop | +0.12  (-0.08, 0.32) | +0.14  (-0.05, 0.32) | -0.13  (-0.34, 0.07) |
| From saturated fat | +0.01  (-0.21, 0.23) | -0.15  (-0.43, 0.13) | Drop | -0.02  (-0.32, 0.27) | **-0.29**  **(-0.49, -0.09)** |
| From monounsaturated fat | -0.01  (-0.20, 0.19) | -0.17  (-0.41, 0.08) | -0.05  (-0.33, 0.22) | Drop | **-0.31**  **(-0.60, -0.01)** |
| From polyunsaturated fat | **+0.58**  **(0.21, 0.96)** | **+0.42**  **(0.02, 0.83)** | **+0.53**  **(0.21, 0.86)** | **+0.55**  **(0.12, 0.98)** | Drop |
| **Sleep latency, minutes** | | | | | |
| From protein | Drop | -0.44  (-1.56, 0.69) | +0.45  (-1.00, 1.91) | **+2.35**  **(0.94, 3.76)** | **-4.38**  **(-6.20, -2.55)** |
| From carbohydrate | +0.44  (-0.73, 1.60) | Drop | +0.93 (-1.29, 3.14) | **+2.82**  **(0.77, 4.88)** | **-3.90**  **(-6.15, -1.65)** |
| From saturated fat | -0.92  (-3.35, 1.51) | -1.31  (-4.39, 1.77) | Drop | +1.47  (-1.76, 4.70) | **-5.25**  **(-7.49, -3.01)** |
| From monounsaturated fat | **-3.70**  **(-5.85, -1.54)** | **-4.09**  **(-6.78, -1.41)** | **-3.21**  **(-6.22, -0.19)** | Drop | **-8.03**  **(-11.27, -4.79)** |
| From polyunsaturated fat | **+10.34**  **(6.22, 14.46)** | **+9.94**  **(5.49, 14.40)** | **+10.83**  **(7.25, 14.41)** | **+12.73**  **(7.96, 17.50)** | Drop |
| **% Wakefulness after sleep onset, %** | | | | | |
| From protein | Drop | +0.15  (-0.51, 0.80) | -0.64  (-1.49, 0.20) | **+1.19**  **(0.36, 2.01)** | **-1.80**  **(-2.86, -0.73)** |
| From carbohydrate | -0.18  (-0.85, 0.50) | Drop | -0.82  (-2.11, 0.48) | +1.01  (-0.19, 2.21) | **-1.97**  **(-3.28, -0.66)** |
| From saturated fat | +1.07  (-0.34, 2.49) | +1.22  (-0.57, 3.02) | Drop | **+2.26**  **(0.38, 4.14)** | -0.72  (-2.02, 0.59) |
| From monounsaturated fat | **-1.81**  **(-3.07, -0.56)** | **-1.66**  **(-3.23, -0.10)** | **-2.45**  **(-4.21, -0.70)** | Drop | **-3.60**  **(-5.49, -1.72)** |
| From polyunsaturated fat | **+4.34**  **(1.93, 6.74)** | **+4.49**  **(1.89, 7.08)** | **+3.70**  **(1.61, 5.78)** | **+5.52**  **(2.74, 8.31)** | Drop |

Note: Data are presented as non-standardized coefficients with 95% confidence intervals. The results in bold are significant (*P* < .05). All estimates have been adjusted for age, sex, body mass index.
